# Using Genomic and Traditional Epidemiologic Approaches to Define Complex Transmission Pathways of *Klebsiella pneumoniae* Infection in a Neonatal Unit in Botswana, 2022–2023

**DOI:** 10.1101/2025.11.06.25339637

**Authors:** Jonathan Strysko, Weiming Hu, Kagiso Mochankana, Janet John-Thubuka, Tshiamo Zankere, Boingotlo Gopolang, Erin Theiller, Steven M. Jones, Chimwemwe Viola Tembo, Tlhalefo Dudu Ntereke, Teresia Gatonye, Kwana Lechiile, Tapoloso Keatholetswe, Colleen Bianco, Susan E Coffin, Carolyn McGann, Kyle Bittinger, Ebbing Lautenbach, Naledi Mannathoko, Margaret Mokomane, Mosepele Mosepele, Melissa Richard-Greenblatt, Britt Nakstad, David M. Goldfarb, Paul Planet, Ahmed M. Moustafa

## Abstract

**Background:** *Klebsiella pneumoniae* (*Kpn*) is a major cause of infant mortality worldwide, with most transmission occurring among hospitalized neonates in low-and middle-income countries where infections caused by multidrug-resistant *Kpn* (MDR-*Kpn*) are increasingly common. We hypothesized that integrating laboratory surveillance for neonatal colonization and infection, real-time epidemiologic investigations, and whole-genome sequencing (WGS) could identify transmission pathways to guide targeted infection prevention and control (IPC) strategies.

**Methods and Findings:** We conducted *Kpn* surveillance in a 36-bed neonatal unit in Botswana over 12 months (2022–2023). WGS was performed on *Kpn* isolates from bloodstream infections (BSIs), and MDR-*Kpn* isolates collected from environmental sampling during outbreaks and twice-monthly colonization screenings (skin and perirectal swabs) using culture media selective for MDR-*Kpn* (CHROMagar Extended-spectrum beta-lactamase [ESBL]/SuperCarba). WGS data were analyzed using multilocus sequence typing (MLST), pangenome and reference-based single-nucleotide polymorphism (SNP) analyses, and Bayesian phylogenetics.

We identified 55 *Kpn* BSIs during the 12-month surveillance period and the median prevalence of MDR-*Kpn* colonization was 28%. *Kpn* was recovered from multi-use intravenous (IV) fluid bags during a *Kpn* outbreak (41 BSIs, 10 deaths), which was controlled by implementing a 24-hour discard policy for IV medications. Among 270 *Kpn* isolates available (28 BSI, 232 colonizing, 10 environmental [six IV fluid, four sink drain]), WGS confirmed over half of BSI genomes (n=17) were ST1414, a clone susceptible to third-generation cephalosporins not detected during MDR-*Kpn* colonization screening, but closely related (<25 SNPs) to six *Kpn* isolates from contaminated IV fluids.

**Conclusions:** This study reinforces the value of integrating WGS with real-time epidemiologic investigations to understand transmission dynamics and guide IPC. Colonization surveillance focused solely on MDR-*Kpn* may overlook drug-susceptible but outbreak-prone strains.

## Introduction

*Klebsiella pneumoniae (Kpn)* is a major cause of neonatal sepsis and sepsis-related mortality, particularly in low-and middle-income countries (LMICs), where it has been implicated in up to 25% of deaths among children under two years of age.^1–3^ In neonates, most *Kpn* infections occur in the context of hospital transmission, highlighting the critical role of infection prevention and control (IPC) measures in curbing its spread.^4–7^ In Botswana, where this study was carried out, bloodstream infection (BSI) among hospitalized neonates is associated with 30% mortality and *Kpn*—especially multidrug-resistant strains—has been shown to be the leading cause.^8^

Whole-genome sequencing (WGS) has emerged as a powerful tool in *Kpn* outbreak investigations, enabling identification of clonal clusters, multilocus sequence types (MLST), antimicrobial resistance genes (ARGs), virulence factors, and K and O capsular serotypes.^6, 9^ WGS may identify clonal outbreaks that may be missed by traditional epidemiologic methods; this is particularly useful when transmission patterns are complex.^10^ The potential impact of WGS methodology on outbreak investigations is especially important in LMIC neonatal care settings, where *Kpn* colonization is high, environmental contamination is common, and IPC practices are often constrained by resource limitations. ^11–13^ Enhanced understanding of *Kpn* reservoirs and routes of transmission within neonatal units could inform targeted remediation strategies to prevent infections in this vulnerable population.

The characterization of *Kpn* isolates from LMICs remains limited, primarily due to constrained diagnostic capacity, including limited availability of culture and especially molecular tools such as WGS. Although prior studies in LMICs have provided compelling genomic evidence of sustained nosocomial transmission of *Kpn*, many have relied on retrospective data sets or infrequent point-prevalence surveys, limiting their ability to pinpoint specific sources.^4, 6^ Others have shown that integrating WGS data with prospective epidemiologic investigations and targeted environmental sampling can elucidate outbreak dynamics.^14–17^ Nevertheless, the transmission pathways contributing to neonatal infections, particularly those involving patient-to-patient and environment-mediated spread, remain poorly understood.^15, 16, 18^ Disentangling these routes requires prospective surveillance of patient colonization and the healthcare environment, alongside the integration of genomic and epidemiologic data.

In this study, we integrated genomic data with prospective epidemiologic investigations to examine *Kpn* transmission in a neonatal unit in Botswana. By analyzing temporally-linked clinical, colonizing, and environmental isolates alongside real-time epidemiologic observations, we sought to identify complex *Kpn* transmission dynamics. Our report demonstrates how WGS, when embedded within a responsive surveillance framework, can inform targeted and effective IPC interventions for outbreak responses in LMICs.

## Methods Study Setting

This study was conducted in a 36-bed neonatal intensive care unit (NICU) within a 530-bed public tertiary referral hospital in Botswana where over 7,000 deliveries occur annually. Common diagnoses in this NICU include prematurity-related complications, hypoxia-related injuries, and sepsis, with multidrug-resistant *Kpn* (MDR-*Kpn*) being the leading cause of BSIs.^8^ Care includes oxygen support, mechanical ventilation, cardio-respiratory monitoring, enteral and parenteral hydration and nutrition, thermoregulation, transfusion, post-surgical care, phototherapy, and fluid/electrolyte management. Due to shortages and routine practices aimed at conserving resources, some medication vials and intravenous (IV) solutions are shared among patients. The hospital has a dedicated IPC program, including two full-time infection prevention nurses. Access to soap, water, and alcohol-based hand sanitizer is generally reliable in the neonatal unit; however, personal protective equipment such as gloves and gowns is frequently out of stock.

### Patient Colonization Isolates

Aggregate colonization prevalence with MDR-*Kpn* was estimated through twice-monthly point prevalence surveys conducted from November 2022 to November 2023. Periumbilical skin and perirectal swabs were collected from all inpatients, including those previously determined to be colonized on preceding surveys. Swabs were processed within 24 hours using chromogenic media (CHROMagar ESBL and SuperCarba, Paris, France) to detect extended-spectrum cephalosporin-resistant and carbapenem-resistant *Kpn,* collectively defined as MDR-*Kpn*. A patient was considered colonized if *Kpn* growth was detected on either swab, identified presumptively through visual growth on selective media, and later confirmed by WGS.

Individual patient identifiers were not collected during these surveys, precluding linkage between individual colonization and BSI outcomes.

### Environmental Isolates

Environmental sampling for MDR-*Kpn* was performed during outbreak periods using nylon flocked swabs on high-touch surfaces, sink drains, medical equipment, and the hands of healthcare workers and mothers using a previously described low-cost technique.^12^ Swab samples were processed similarly to colonization swabs and MDR-*Kpn* species were identified presumptively through visual growth on the selective media, and later confirmed by WGS. Five milliliter samples of IV fluids and medications were inoculated into pediatric blood culture bottles for incubation. Presumptive MDR-*Kpn* species identified from IV fluids and medications were identified manually via Gram stain and colony morphology, followed by sub-culturing and WGS species confirmation.

### Blood Culture Isolates

Neonatal blood cultures were collected from November 2, 2022 to November 7, 2023 by hospital staff based on clinical indications and incubated at the hospital microbiology laboratory using an automated system (BACT/ALERT, BioMérieux). There were no blood culture bottle shortages during this period, and patients were not required to pay for blood cultures, as is the case in some hospitals. Isolates were identified manually via Gram stain and colony morphology, followed by sub-culturing. Antimicrobial susceptibility testing (AST) was conducted using manual disc diffusion testing; however, due to shortages of antibiotic discs for AST, all BSI isolates in this analysis underwent automated identification and AST (VITEK 2, BioMérieux) using Clinical & Laboratory Standards Institute (CLSI) minimum inhibitory concentration (MIC) breakpoints.^19^ Isolates were classified as non-susceptible if they demonstrated intermediate susceptibility or resistance according to CLSI standards. *Kpn* BSI isolates identified by WGS as the same ST and recovered from the same patient within a 14-day period were considered duplicates; only the first isolate was included.

### Epidemiologic Methods

The neonatal unit does not have a standard outbreak threshold, but clinicians consider two or more cases of gram-negative BSI within one week as a sentinel event. When this occurs, the hospital’s IPC team is prompted to start a line list to document cases and exposures. During the third month of this surveillance period, an increased number of *Kpn* BSI were noted. Confirmed cases were defined as laboratory-confirmed BSI caused by *Kpn* occurring within one month prior to the observed increase in cases. The line list recorded patient age at symptom onset, sex, ward location, culture results, AST results, and outcomes (discharged/died). Regular outbreak response meetings attended by ward clinicians and IPC team members were held to review trends, generate hypotheses, and implement interim control measures.

### Genomic Analysis

Single colonies of suspected *Kpn* isolates from clinical, colonizing, and environmental samples were harvested from culture plates and stored as glycerol stocks at-80°C for genomic analysis. DNA was extracted using the Illumina Direct colony extraction protocol and quantified using the Quant-iT PicoGreen dsDNA assay kit (Thermo Fisher Scientific). Shotgun libraries were prepared using the Illumina DNA Prep kit with unique dual indices. Libraries were quantified, and samples yielding <1 ng/μL were re-prepared. Equal volumes from each library were pooled and initially sequenced on a MiSeq Nano run to guide final pooling before sequencing on a NovaSeq 6000 (2×150 base pairs). Negative controls and a mock community (*Vibrio campbellii* and Lambda phage DNA) were included to monitor contamination and sequencing performance.

Raw sequences were passed through Sunbeam v4.7.0 pipeline for quality control prior to assembly.^20^ Within the pipeline, Illumina PhiX adapters, low sequence complexity sequences and human sequences were removed. The SPAdes v3.15.5 isolate model was used to assemble high quality reads.^21^ The quality of the assembled contigs was assessed by CheckM v1 and Mash.^22, 23^ Assemblies potentially contaminated by other genomes (more than 100 shared hashes with other species out of 1000 total hashes, or completeness score greater than 95% and contamination score greater than 5% based on CheckM) went through the avamb pipeline to identify single genomes.^24^ The samples identified as having high quality genomes (less than 10% shared hashes with other species and completeness greater than 95% and contamination less than 5% based on CheckM) from both steps were annotated using Bakta v1.9.1.^25^

Panaroo v1.5.0 was used to build pangenome analysis alignment, with mafft as the aligner.^26, 27^ The pairwise single-nucleotide polymorphism (SNP) distance between samples were calculated using snip-dists based on the pangenome core genome alignment.^28^ A phylogenetic tree of the pangenome alignment (3,263,164 base pairs) was built using IQ-TREE v2.3.3 with ‘GTR+F+I+R8’ as the DNA model with 1000 bootstrap replicates.^29^ For each sequence type (ST), the best-quality genome among the study isolates was selected as the reference. SNP distances were then calculated using Snippy relative to this reference for all other study isolates of the same ST as well as for closely related genomes of the specific ST. Closely related external public genomes were chosen using the *top genome* feature in WhatsGNU^30–32^. Accessions for selected external public genomes are available.^33^ For each ST, the evolution time of the phylogenetic tree was calculated using Bayesian evolutionary analysis using BEAST v 2.7.7^34^ Specifically, the above mentioned SNP alignment FASTA file was imported into Beauti, the tip date was relative to the most recent isolate which was set as 0, HKY was used as the subset model, strict clock was used and the clock rate distribution was set as log-normal. Coalescent constant population was used as priors, and the Markov chain Monte Carlo (MCMC) chain length was set at 10e8. After inspection of the MCMC traces and the effective sample size (>200) values of each run, 10% of the first posterior samples were removed as a burn-in. The chronogram was plotted on the basis of the maximum clade credibility tree using the TreeAnnotator program from the BEAST package.

*Kpn* virulence factors and ARGs were identified using kleborate v3.^35^ All figures, including phylogenetic trees, were generated in R v4.4.2 using the following packages: ggplot2, ape, ggtree, treeio, ggtreeExtra, patchwork, ggstar, aplot, tidytree.^36^ Static *Kpn* clusters are often defined as strains sharing a recent common ancestor with ≤21 pairwise SNP differences.^37^ Thus, heatmaps were stratified with scale ranges from 0–25 SNPs (suggesting very close genetic relatedness, indicative of recent transmission) to 201–8000 SNPs (suggesting distant evolutionary divergence).

To infer the most likely source of the outbreak, we performed ancestral state reconstruction using PastML v1.9.51, a parsimony-based algorithm for mapping discrete traits onto the phylogenetic tree (see Supplementary material).^38^ The time-scaled BEAST phylogeny of ST-matched isolates was used as input, and isolate source type (clinical and environmental) was assigned as a categorical trait. Ancestral states were inferred under the Maximum Parsimony Accelerated Transformation model with default parameters. The resulting annotated tree was visualized to identify the most probable origin and dissemination route of the outbreak strain within the NICU.

### Ethical Review

This study was approved by the Institutional Review Boards at the University of Botswana (UBR/RES/IRB/BIO/305), the Health Research and Development Committee at Botswana’s Ministry of Health (2022-HPDME18/13/1), and the healthcare facility where this study was carried out (2022-2/2A (7)/201). Because this study relied on surveillance data and did not collect identifiable information, written patient consent was waived. All methods adhered to the principles of the Declaration of Helsinki.

## Data availability

Sample-level data are described in **Supplementary Table 1.** Raw whole-genome sequence data were deposited under the following BioProject: PRJNA1271068 and PRJNA1291254. Raw tree and heatmap files for main and supplementary figures are available for download.^33^

## Results

During the 12-month surveillance period, a total of 96 BSIs were detected among 1093 infants admitted to the study NICU, 55 of them due to presumed *Kpn* (**Figure 1A**). Among 1562 skin and perirectal colonization screening samples taken during the 12-month study period, the median ward prevalence of skin or perirectal colonization with MDR-*Kpn* (species confirmed by WGS) was 28% (IQR: 21%-36%) (**Figure 1B**).

**Figure 1.**
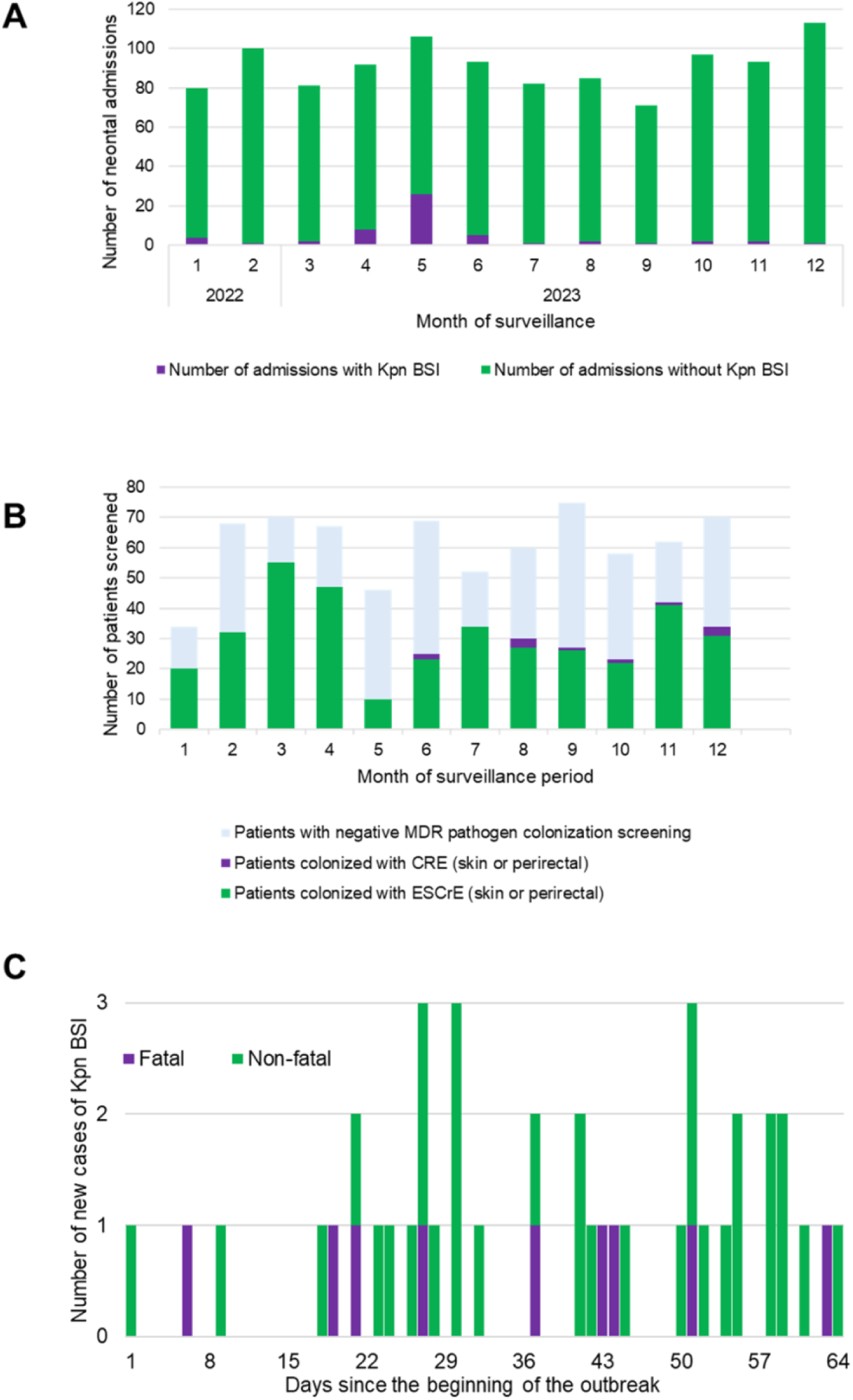
*Klebsiella pneumoniae* (*Kpn*) colonization and bloodstream infection in a neonatal unit in Gaborone, Botswana (2022–2023). **A.** Monthly number of admissions with and without *Kpn* BSIs; **B.** Prevalence of skin or perirectal colonization of multidrug-resistant Enterobacterales (presumptive extended-spectrum cephalosporin-resistant *Enterobacterales* [ESCrE] or carbapenem-resistant Enterobacterales [CRE]) detected during twice-monthly ward screening. **C.** Epidemic curve of *Kpn* bloodstream infections (BSI), stratified by fatal vs. non-fatal outcome detected during outbreak period, 2023.

### Epidemiologic findings during *Kpn* outbreak

During this surveillance period, the hospital’s IPC team was alerted by NICU clinicians to an unexpected rise in cases of *Kpn* BSI. A line list was initiated to track confirmed and suspected cases. Over a nine-week period, a total of 41 confirmed *Kpn* BSI cases were identified, with 10 fatalities among the affected patients (**Figure 1C**, **Table 1)**.

**Table 1.** Epidemiology and outcomes for patients with neonatal bloodstream infection with *Klebsiella pneumoniae*, alongside sequence type (performed later through WGS) 2023, Gaborone, Botswana; (GA=Gestational age)

| Case | Onset age (range in days) | GA (range in weeks) | Outcome | Sequence Type |
| --- | --- | --- | --- | --- |
| 1 | 0-5 | 36-40 | Discharged |  |
| 2 | 11-15 | 31-35 | **Died** |  |
| 3 | 21-25 |  | Discharged | ST4 |
| 4 | 0-5 |  | Discharged |  |
| 5 | 11-15 | 31-35 | **Died** |  |
| 6 | 6-10 | 26-30 | **Died** | ST4083 |
| 7 | 6-10 | 31-35 | Discharged |  |
| 8 | 6-10 | 31-35 | Discharged |  |
| 9 | 0-5 | 26-30 | Discharged |  |
| 10 | 0-5 |  | Discharged |  |
| 11 | 16-20 |  | Discharged | ST1414 |
| 12 | 0-5 | 31-35 | **Died** |  |
| 13 | 0-5 |  | Discharged |  |
| 14 |  | 36-40 | Discharged |  |
| 15 | 0-5 | 31-35 | Discharged |  |
| 16 | 6-10 | 26-30 | Discharged | ST1414 |
| 17 | 0-5 | 26-30 | Discharged |  |
| 18 | 6-10 | 26-30 | **Died** | ST17 |
| 19 |  |  |  | ST1414 |
| 20 | 0-5 | 36-40 | Discharged | ST1414 |
| 21 | 6-10 |  |  | ST1414 |
| 22 | 0-5 | 26-30 | **Died** | ST1414 |
| 23 | 0-5 | 36-40 | Discharged |  |
| 24 | 0-5 | 26-30 | **Died** | ST1414 |
| 25 | 0-5 | 36-40 | Discharged |  |
| 26 | 11-15 | 26-30 | Discharged |  |
| 27 | 0-5 | 26-30 | Discharged |  |
| 28 | 11-15 | 21-25 | **Died** |  |
| 29 | 0-5 | 31-35 | Discharged |  |
| 30 | 0-5 | 36-40 | Discharged | ST1414 |
| 31 | 0-5 | 31-35 | Discharged | ST1414 |
| 32 | 0-5 | 31-35 | Discharged |  |
| 33 | 0-5 |  | Discharged |  |
| 34 | 21-25 | 31-35 | Discharged |  |
| 35 | 0-5 | 31-35 | Discharged | ST1414 |
| 36 | 0-5 | 31-35 | Discharged |  |
| 37 | 0-5 |  | Discharged | ST1414 |
| 38 | 6-10 |  | Discharged | ST1414 |
| 39 |  | 31-35 | **Died** | ST1414 |
| 40 | 0-5 | 26-30 | **Died** | ST1414 |
| 41 | 0-5 | 31-35 | Discharged | ST1414 |

A point prevalence survey of environmental samples conducted during the outbreak included 60 samples collected from surfaces and medical equipment and plated on selective agars as described above.

Presumptive MDR-*Kpn* species were detected in four sampled sink drains. All other samples, including those taken from bed rails, suction tubing, suction canisters, feeding utensil storage buckets, and 17 samples from the hands of healthcare workers and mothers, showed no growth of MDR*-Kpn*.

On week eight of the outbreak, noting low prevalence of MDR-*Kpn* colonization unit-wide, early age of onset (four days) of patients, and a high likelihood of having received IV medications soon after birth, the IPC team considered possible vehicles that could lead to direct inoculation of *Kpn* without prior colonization (**Table 2)**. This prompted sampling of IV fluids and medications. Six dextrose-containing IV fluid bags which had been opened and were in use were found to be contaminated with presumed *Kpn*. In contrast, unopened IV fluid bags showed no growth. Given these observations, interim outbreak measures were instituted (**Table 3**). A strict policy to discard all IV fluids and medications within 24 hours of opening was implemented, effectively ending the outbreak.

**Table 2.** Real-time epidemiologic observations made during the outbreak of *Klebsiella pneumoniae* (*Kpn*) bloodstream infections, 2023, Gaborone, Botswana.

| Observation | Implications |
| --- | --- |
| 1. The median postnatal age of onset was low (four days) with 30% (n=11) of infants having a positive blood culture within in the first two days of life. | Early exposures either in the neonatal unit or the labor ward were likely driving transmission. |
| 2. The majority of cases occurred among preterm patients (median estimated gestational age of 32 weeks) who were first admitted to the premature or intensive care cubicles | Shared exposures in the premature and NICU wards, such as medications, staff, and equipment, were likely driving transmission. In the premature ward, patients receive supportive care with early IV hydration and medication (e.g. antibiotics), enteral feeding and medication (e.g. caffeine) with nasogastric tubes, humidified oxygen supplementation, and thermoregulation with radiant warmers and incubators. |
| 3. Environmental sampling of surfaces and equipment in the unit did not identify *Kpn*. | Transmission was either not occurring through fomites or it was occurring through un-sampled fomites. |
| 4. Hand cultures from staff and families did not grow *Kpn*. | Transmission was either not occurring from contaminated hands, or staff and mothers performed hand hygiene prior to sampling, or transmission was occurring from contaminated hands of staff/families who were not included in the sampling event. |
| 5. Ward prevalence of MDR-*Kpn* colonization reached a nadir in the middle of the outbreak. | Patients were likely not pre-colonized with *Kpn* prior to becoming bacteremic, or colonization screening was not detecting drug-susceptible strains. |
| 6. Several patients re-cultured *Kpn* despite several days of being on appropriate antibiotic therapy. | Patients were either missing doses of antibiotics, had internal abscesses which antibiotics could not penetrate, or were being re-infected through a contaminated vehicle. |
| 7. Three cases of *Enterobacter* BSI occurred during the outbreak, including two cases in patients co-infected with *Kpn* BSI. | Co-occurrence of *Enterobacter* BSI during an ongoing *Kpn* outbreak, including cases of co-infection with *Kpn* BSI suggested common transmission pathways despite being different organisms. |
| 8. Samples from six opened multi-use IV fluids bags cultured *Kpn*. | Contaminated IV fluid bags were likely a major vehicle for *Kpn* transmission. |

**Table 3.** Outbreak response measures made during the outbreak of Klebsiella pneumoniae bloodstream infections, 2023, Gaborone, Botswana.

| Outbreak response measures |  |
| --- | --- |
| 1. | Patients with suspected and confirmed BSI placed on contact precautions and cohorted in isolation rooms. |
| 2. | Cleaning and disinfection of surfaces and medical equipment intensified, and rooms fumigated with sodium hypochlorite-based disinfection product. |
| 3. | Hand hygiene education intensified for staff and families. |
| 4. | Feeding utensil cleaning and disinfection method overhauled, shifting from communal chemical disinfection to individualized thermal disinfection using microwave steam bags. |
| 5. | All infants >24 hours old and weighing >1 kg cleansed with aqueous 2% chlorhexidine gluconate twice weekly. |
| 6. | All opened IV fluid bags discarded, and a unit policy was instituted requiring the labelling of all IV fluid bags upon opening and discarding within 24 hours after opening. |

### Genomic analysis

#### Phylogenetic analysis

Only 32 of the 55 BSI isolates were submitted for sequencing (23 discarded prior to preservation), of which 88% (n=28) were confirmed as *Kpn* by WGS. Among 370 colonization isolates, and 19 environmental isolates from sinks submitted for sequencing, 63% (n=232), and 21% (n=4) were confirmed as *Kpn* respectively. The lack of specificity of the ESBL chromogenic media is largely due to *Kpn* colony appearance (metallic blue) being the same for *Enterobacter* spp., *Citrobacter* spp., and all other *Klebsiella* spp. All six isolates from IV fluids were confirmed as *Kpn* **(Figure 2A)**.

**Figure 2.**
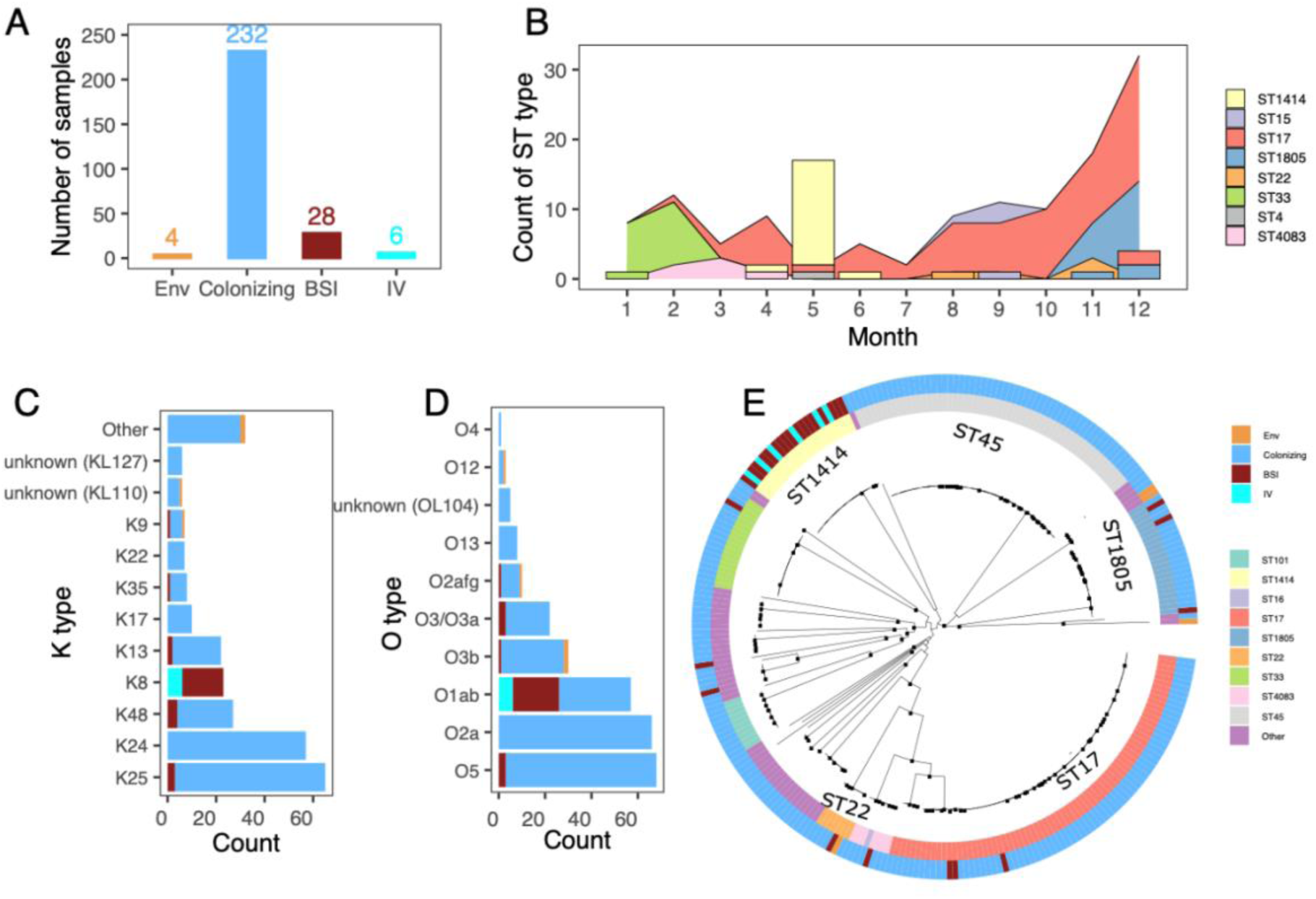
Molecular Epidemiology and Genomic Diversity of *Klebsiella pneumoniae* (*Kpn*) Isolates Collected During Surveillance in a Neonatal Unit in Gaborone, Botswana (2022–2023). A. Distribution of confirmed Kpn samples by source: environmental (Env), colonizing, bloodstream infections (BSI), and intravenous (IV) fluid-associated isolates (n = 270 total). **B.** Monthly prevalence of Kpn colonization and incidence of *Kpn* BSI. Colors represent different sequence types (STs), curves represent colonization prevalence and bars represent BSI incidence. **C-D.** Histogram of capsule (K) and lipopolysaccharide (O) antigen types identified among, stratified by source (BSI, colonizing, environmental, and contaminated IV fluid isolates). **E.** Maximum likelihood phylogenetic tree based on core genome alignment of all study Kpn colonizing, BSI and environmental isolates. The inner ring is color-coded by ST, and the outer ring by isolate source. Bootstrap values above 75 are shown on the branches as black squares.

The maximum likelihood phylogenetic tree of the 270 *Kpn* isolates highlights the extensive diversity of *Kpn* lineages circulating within the NICU, with multiple distinct clades representing both colonizing and invasive isolates, each encompassing diverse sequence types (STs) and K and O antigen types (**Figure 2B, C, D and E**). This genomic heterogeneity suggests a complex transmission ecology in the unit, with both sporadic and outbreak-related dynamics. WGS revealed substantial diversity among MDR-*Kpn* strains, identifying 28 distinct STs among colonization isolates, with ST17 and ST45 accounting for the most common STs among colonizing MDR-*Kpn* isolates. ST45, despite its dominance as a colonizing strain, was never associated with invasive disease during the study period (**Supplementary Figure 1 and Supplementary Table 2**).

Among BSI isolates, eight STs were identified: ST4 (n=1), ST15 (n=1), ST17 (n=3), ST22 (n=1), ST33 (n=1), ST1414 (n=17), ST1805 (n=3), and ST4083 (n=1) (**Figure 2B**, **2E**). All but two of these STs (ST1414 and ST4) were also detected in colonization samples (**Figure 2B**). **Figure 3** depicts the estimated the time to most recent common ancestor (tMRCA) for each BSI-associated ST, with the x-axis representing time, ranging from approximately 2022 to 2023 and suggests varying divergence timelines, reflecting distinct evolutionary trajectories. ST1414 was identified in 17 BSI isolates, all occurring during the 9-week outbreak period and in all six contaminated IV fluid bags, but it was not detected during colonization surveillance. Since all ST1414 BSI isolates were found to be phenotypically susceptible to third-generation cephalosporins, (**Supplementary Table 2**), its absence from colonization screening may either reflect direct inoculation without prior colonization (through contaminated IV fluid administration) or suppression by culture media which was selective for MDR-*Kpn.* Phylogenetic, temporal, and ancestral state reconstruction analyses indicate that the ST1414 clone was a recent introduction to the unit, with the estimated tMRCA occurring approximately three months prior to its initial clinical detection in early 2023 (**Figure 4** and **Supplementary Figure 3**). While undetected ST1414 colonization may have contributed to early transmission, ancestral state reconstruction using a parsimony-based approach (PastML) suggests that contaminated IV fluid was the source of the outbreak. Although all IV and BSI isolates were closely related, the IV isolates did not form a single clade. Instead, they appeared across multiple subclades, each intermixed with bloodstream infection isolates. This pattern suggests repeated introductions of the outbreak strain via contaminated IV fluids, rather than a single contamination event (**Figure 4B)**.

**Figure 3.**
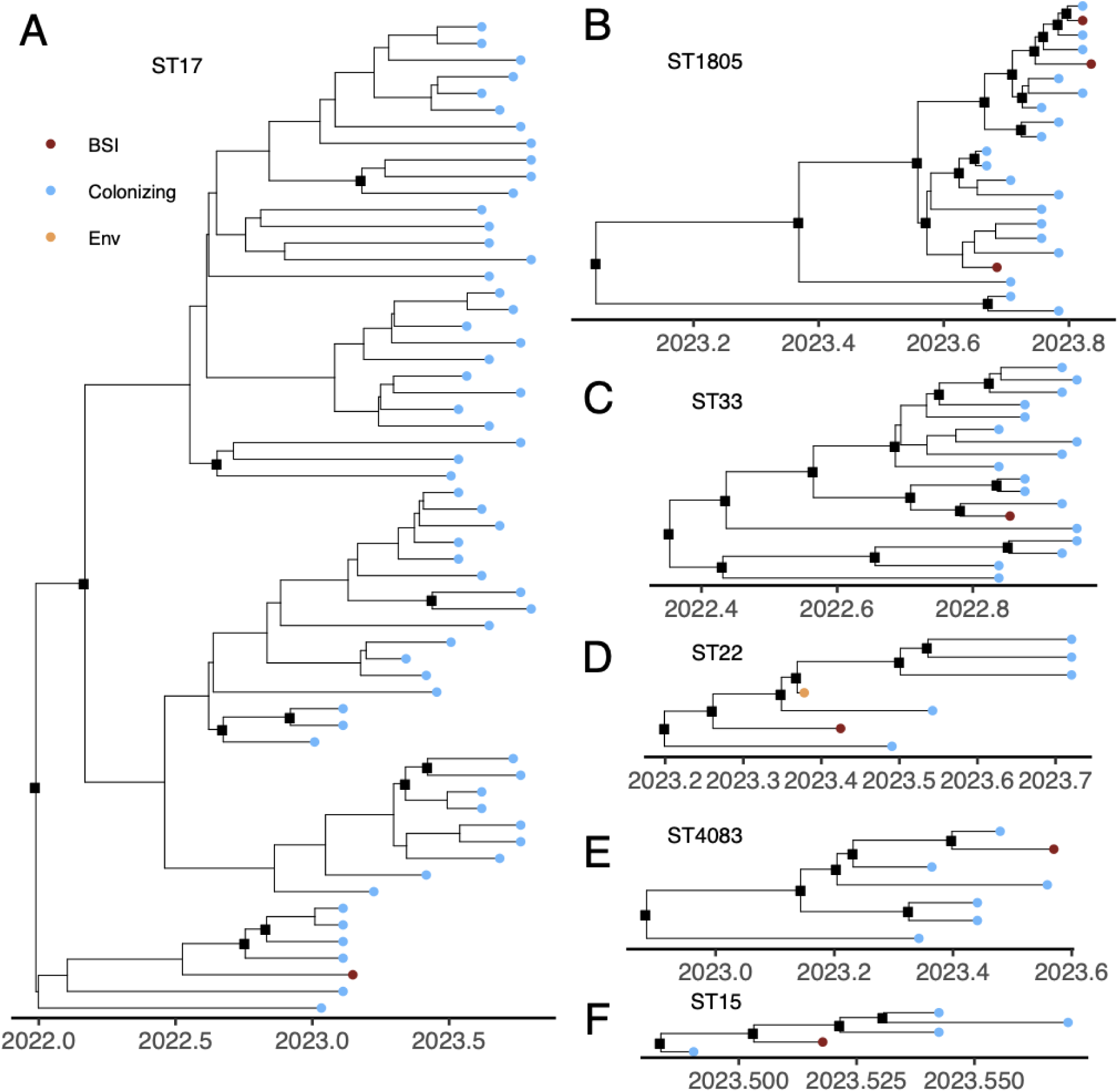
Time-scaled BEAST phylogenies of *Klebsiella pneumoniae* sequence types associated with bloodstream infections. A-F. BEAST trees are shown for all *K. pneumoniae* sequence types (STs) identified in bloodstream infection (BSI) cases, including **A.** ST17, **B.** ST1805, **C.** ST33, **D.** ST22, **E.** ST4083, and **F.** ST15. Tip colors indicate isolate source (e.g., blue for colonization, red BSI, orange environmental), and posterior branch supports above 0.75 are indicated where relevant with black square.

**Figure 4.**
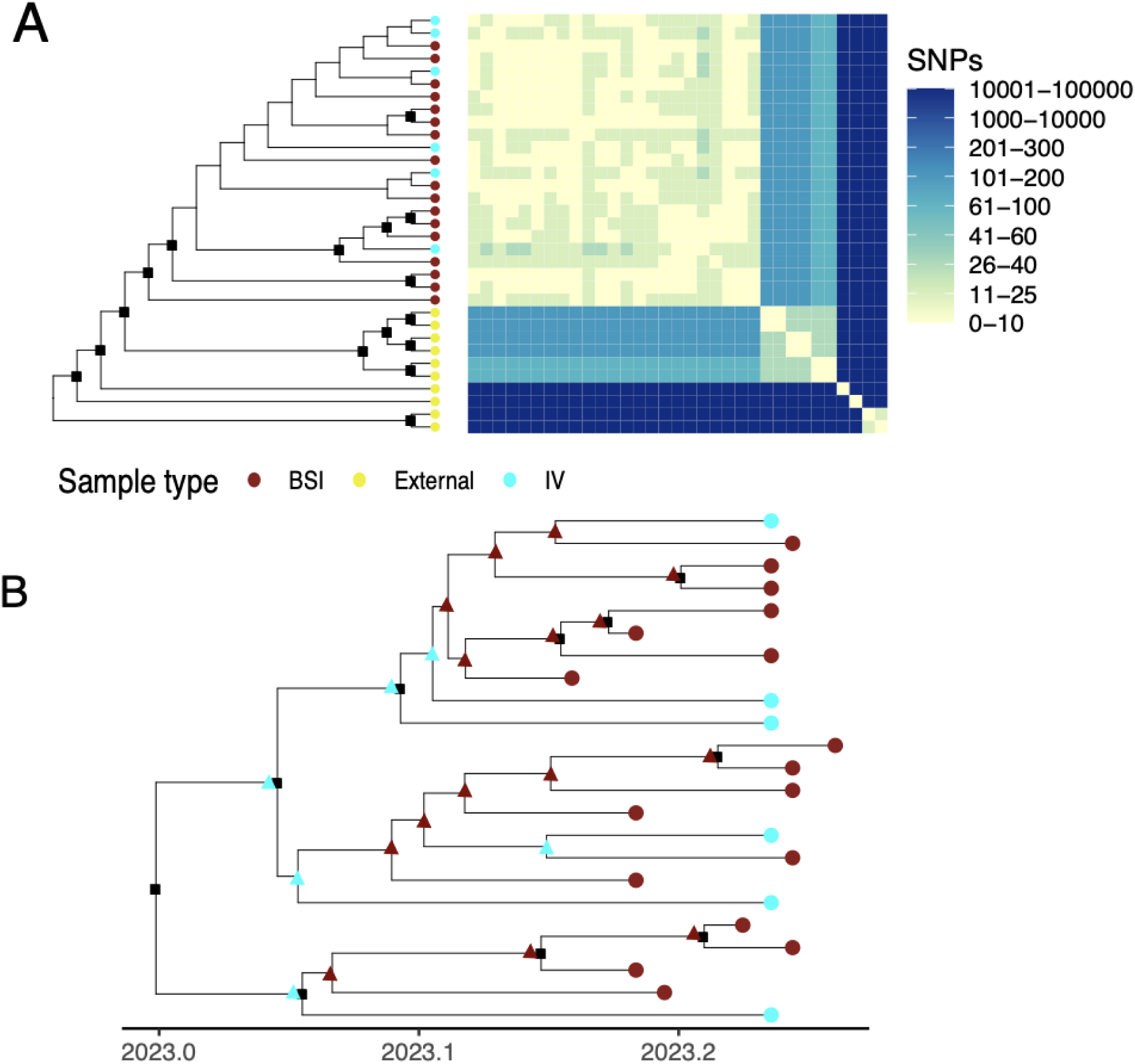
ST1414 *Klebsiella pneumoniae* outbreak phylogeny with ancestral state reconstruction highlights intravenous (IV) fluid bag as a transmission route. A. Maximum likelihood phylogenetic tree of ST1414 isolates shown as a cladogram based on alignment to the best quality genome ENV.NNU.28.dextrose.dup within the ST using Snippy and a heatmap produced using SNP-dist. Bootstrap values above 75 are shown on the branches as black squares. **B**. BEAST tree of ST1414 isolates associated with the outbreak. Tip colors indicate isolate source (red for bloodstream infection [BSI] isolates, cyan for IV fluid isolates, and yellow for external public genomes), and posterior branch supports above 0.75 are marked with a black square where relevant. Ancestral state reconstruction using a parsimony-based approach. The red triangle represents the inferred ancestral node to be BSI-associated which means that the internal node more closely related to descendant isolates from BSI isolates and purple triangle represents the Inferred ancestral node to be IV fluid-associated which means that the internal node more closely related to descendant isolates from IV fluid isolates. Ancestral state reconstruction suggests that contaminated IV fluid played a major role in the outbreak and as the most likely route of transmission; IV fluid isolates were distributed across the ST1414 outbreak clade and yet were closely related to all 17 ST1414 BSI isolates.

Detection of ST22 *Kpn* from a sink drain sample preceded its identification in a blood culture by several weeks and in five colonization samples by several months (**Figure 3D** and **Supplementary Figure 4**). ST17 was consistently detected in colonization samples across the study period and was associated with two temporally related and one unrelated infection (**Figure 3C**).

The persistent detection of ST17 and ST45 along with their phylogenies suggest that they had likely been circulating locally for several years with more recent diversification leading to the emergence of hospital-specific strains **(Figure 3A, Supplementary Figures 1** and 5 and **Supplementary Table**). In contrast, ST22 exhibited a more recent tMRCA, with divergence occurring around 2023, suggesting a relatively recent introduction of this strain (**Figure 3D**). Similarly, ST1414 demonstrated divergence from a common ancestor around 2023, further emphasizing the recent emergence of this ST within the outbreak (**Figure 4**). ST1805 shows a tMRCA around the end of 2022, suggesting that this lineage recently expanded in the hospital environment before BSI detection (**Supplementary Figure 6**).

#### K and O antigen types, virulence factors, and ARGs

K-antigen types were highly specific to and strongly associated with STs (**Supplementary Figures 3, 4, 5, 6, and 7**). For example, ST1414 was exclusively associated with K8, clearly distinguishing it from other lineages. In contrast, O-antigen types were less discriminatory as several STs, including both colonizing and invasive strains, shared the O1ab type, limiting its value for epidemiologic differentiation.

We detected limited virulence factors among the 28 BSI isolates; yersiniabactin, a siderophore associated with enhanced survival, was the only virulence locus identified, present in a subset of isolates with lineages ybt 10 (ICEKp4) and ybt 14 (ICEKp5 or ICEKp12), while aerobactin, salmochelin, colibactin, and rmp-associated genes were absent (**Supplementary Table 1**).

Among 28 BSI isolates, Kleborate analysis revealed the presence of β-lactamase genes, most commonly Sulfhydryl Variable (SHV). Acquired beta lactamase genes included oxacillinase (OXA), temoniera (TEM), and genes encoding for ESBLs, including cefotaximase-Munich (CTX-M)-14 and CTX-15. No carbapenemase genes were detected. Resistance determinants were also identified across other classes, including tet(A) or tet(D) (tetracyclines), sul1/sul2/sul3 (sulfonamides), and mphA (macrolides).

Phenotypic AST was compared with the presence of ARGs in all 28 bloodstream infection (BSI) isolates (**Supplementary Table 2**). Most *Kpn* isolates that were phenotypically non-susceptible to a given antibiotic class (e.g., aminoglycosides, β-lactams) also harbored corresponding ARGs. Despite historical increases in MDR-*Kpn* in this setting, most BSI isolates in this analysis were susceptible to third-generation cephalosporins, and ESBL-associated ARGs (CTX-M-14, CTX-M-15) were detected in only six isolates across five STs (ST15, ST17, ST1805, ST22, and ST33). ARG profiles were more consistent across isolates than phenotypic AST results. For example, despite being <25 SNPs apart and carrying identical ARG profiles, ST1414 displayed variable phenotypic aminoglycoside and beta-lactam susceptibility. This discrepancy likely reflects differences in gene expression, heteroresistance, or assay variability at MIC breakpoints, rather than underlying genomic divergence and highlights that phenotypic susceptibility, sometimes used as a surrogate for strain relatedness during outbreak investigations when ST is unknown, may be unreliable. No phenotypic or genotypic resistance to carbapenems was detected in either BSI, colonization, or environmental *Kpn* isolates.

Together, these results reveal two distinct epidemiological patterns in the NICU: a single, large, point-source outbreak driven by ST1414 linked to contaminated IV fluids, and ongoing sporadic invasive disease caused by diverse colonizing strains.

## Discussion

In this report, we describe how the integration of traditional epidemiologic investigations with WGS data from clinical, colonizing, and environmental isolates elucidated transmission of *Kpn* in a NICU in Botswana. Genomic analyses were essential to differentiate outbreak-related from endemic isolates in this setting where *Kpn* is endemic, while epidemiologic data and real-time observations identified an unsuspected transmission vehicle. Despite the polyclonal nature of BSI cases in this unit, WGS revealed that the majority of temporally clustered BSI were from a unique ST (ST1414). Focused epidemiologic analyses identified exposures common to neonates infected with ST1414. Real-time observations of clinical practice in the unit led to the hypothesis that contaminated bags of IV fluids were the reservoirs of these isolates and that repeated use of fluids drawn from these bags (to reconstitute medications and flush IV catheters) was the probable mechanism of transmission to infected patients. Finally, phylogenetic analyses of isolates from both IV fluid bags and patients established this practice was the driver of this outbreak and led to a practice change which halted the outbreak.

The sudden appearance of ST1414 in both BSI and IV-associated samples, without prior colonization detection, emphasizes the value of longitudinal surveillance in identifying unexpected introductions but also highlights the pitfalls of colonization screening which focuses on multidrug-resistant strains alone. Ancestral state reconstruction using PastML applied to the ST1414 time-scaled phylogeny supported IV fluid as the most probable source, with a tMRCA dating to approximately three months prior to the first recognized BSI case, aligning with the epidemiologic curve. This study also highlights the importance of having blood culture testing available in order to quickly identify potential point source or other outbreaks. Unfortunately, most laboratories in LMIC settings still do not have access to blood culture testing.^39^

Multilocus sequence typing and phylogenetic analysis demonstrated that ST45 and ST17 were endemic in both colonized and infected patients in this neonatal unit, with both lineages detected frequently in colonization samples and occasionally in infections. Both ST45 and ST17 are globally recognized clones known to cause healthcare associated infections.^6, 40^ ST17 was a frequent colonizer throughout the study period, but rarely caused invasive disease, appearing in only a small number of BSI cases. The detection of ST22 *Kpn* from a sink drain sample predated its detection in a blood culture by several weeks, and several months before it was detected in five colonization samples, suggesting the sink might have been a reservoir for a strain that subsequently infected a patient. Sink drains are likely major reservoirs for *Kpn* in neonatal units, seeding the environment through droplet dispersal and leading to colonization of patients and staff.^41^ More research is needed to understand how contaminated sink drains can be remediated and maintained to sustainably prevent contamination with and transmission of nosocomial pathogens.

Contaminated IV fluids have been well-documented as vehicles for neonatal *Kpn* infection, and this study reinforces the importance of medication safety as a critical but often overlooked IPC measure in resource-limited settings.^14, 42^ Medication shortages in such settings often necessitate practices like using multi-dose medication vials or sharing of IV fluids among patients, particularly in neonatal and pediatric wards where weight-based dosing requires small volumes per patient.^43, 44^ The World Health Organization has issued strong recommendations against sharing of IV fluid or solution bags among patients,^45^ but in resource-limited settings prone to medication stockouts, practical contingency measures such as labeling of opened IV fluid bags, discarding them within a set time frame (e.g., 24 hours), and ensuring aseptic techniques when accessing medications can help limit the spread of infection.

The deployment of WGS was key in confirming outbreak sources and transmission patterns in this study. Efforts should prioritize not only enhancing the resolution and routine implementation of colonization surveillance but also integrating real-time genomic tools into outbreak response. Expanding access to WGS in LMICs is essential for timely outbreak source identification, characterization of antimicrobial resistance patterns, and the development of targeted infection prevention strategies. While resource-intensive, WGS provides critical insights in neonatal units where *Kpn* infections are hyperendemic. In such settings, portable and decentralized platforms such as nanopore sequencing may offer a more feasible and cost-effective approach to implementing WGS.^46^

### Limitations

This investigation had several limitations. Colonization screening was anonymized and not linked to individual patients, precluding analysis of direct progression from colonization to infection. The use of selective media biased detection toward resistant strains, preventing the identification of colonization by *Kpn* strains susceptible to third-generation cephalosporins, of which ST1414 was one. Environmental sampling was limited to peri-outbreak point-prevalence surveys, offering only a snapshot of contamination. More frequent or longitudinal sampling could have better characterized reservoirs and timing. Additionally, data on clinical practices, such as IV fluid handling and medication preparation, were not systematically collected, limiting the ability to correlate observed transmission with procedural lapses.

## Conclusions

This investigation highlights the critical importance of integrating real-time epidemiologic observations with genomic surveillance to identify and confirm sources of healthcare-associated outbreaks of *Kpn*. The detection of contaminated IV fluids as the primary vehicle for transmission underscores the need to strengthen IPC practices focused on medication safety, particularly in resource-limited settings. Practical measures such as labeling opened IV fluid bags, discarding them within defined timeframes, and maintaining strict aseptic technique are essential safeguards when single-use medications are not consistently available. WGS, which offered high-resolution insights into transmission dynamics, was greatly enhanced by traditional field-based epidemiology and targeted environmental sampling. This combined approach was essential for interpreting complex transmission patterns and distinguishing endemic colonization from true point-source introductions. Efforts should prioritize improving the resolution and implementation of colonization surveillance, exploring real-time WGS deployment during outbreaks with decentralized nanopore sequencing, and addressing systemic vulnerabilities like medication shortages and weak IPC infrastructure which increase the risk of future outbreaks.

## Data Availability

Sample-level data are described in Supplementary Table 1. Raw whole-genome sequence data were deposited under the following BioProject: PRJNA1271068 and PRJNA1291254. Raw tree and heatmap files for main and supplementary figures are available for download.33

https://zenodo.org/records/17102790

## Acknowledgements

We would like to thank the sequencing and computational genomics teams at the CHOP Microbiome Center (Philadelphia). We thank the clinical and laboratory staff involved in collection and processing of relevant samples and isolates.

## Author contribution statement

**Conceptualization**: Jonathan Strysko, Susan Coffin, Paul Planet and Ahmed Moustafa

**Data Curation**: Jonathan Strysko, Weiming Hu, Teresia Gatonye, Tlhalefo Ntereke, Tshiamo Zankere, Kwana Lechiile.

**Formal Analysis**: Jonathan Strysko, Weiming Hu, Steven M. Jones and Ahmed M. Moustafa.

**Funding Acquisition**: Susan Coffin, Jonathan Strysko, Kyle Bittinger, Ahmed M. Moustafa.

**Investigation**: Kagiso Mochankana, Melissa Richard-Greenblatt, Viola Tembo, Boingotlo Gopolang, Janet John-Thubuka, Tapoloso Keatholetswe, Kyle Bittinger, Colleen Bianco, Carolyn McGann, Paul Planet, Jonathan Strysko, Britt Nakstad, David M. Goldfarb, Naledi Mannathoko, Mosepele Mosepele, Margaret Mokomane, Erin Theiller, Ahmed M. Moustafa.

**Methodology**: Jonathan Strysko, Susan Coffin, Weiming Hu, Paul Planet, Ahmed M. Moustafa

**Resources**: Susan Coffin, Jonathan Strysko, Ebbing Lautenbach, Ahmed M. Moustafa

**Supervision**: Jonathan Strysko, Susan Coffin, Kyle Bittinger, Ebbing Lautenbach, Paul Planet, Ahmed M. Moustafa

**Visualization**: Jonathan Strysko and Weiming Hu.

**Writing – Original Draft Preparation**: Jonathan Strysko and Ahmed M. Moustafa

**Writing – Review & Editing**: All authors.

**Supplementary Table 1.** Sample-level metadata and whole-genome sequencing information for isolates included in this study.

**Supplementary Table 2**. Phenotypic antimicrobial susceptibility testing results (Clinical & Laboratory Standards Institute standards) alongside presence of antimicrobial resistance genes identified for 28 bloodstream infection *Klebsiella pneumoniae* isolates, by antibiotic class and sequence type, Gaborone, Botswana, 2022–2023

---

**Supplementary Figure 1.**
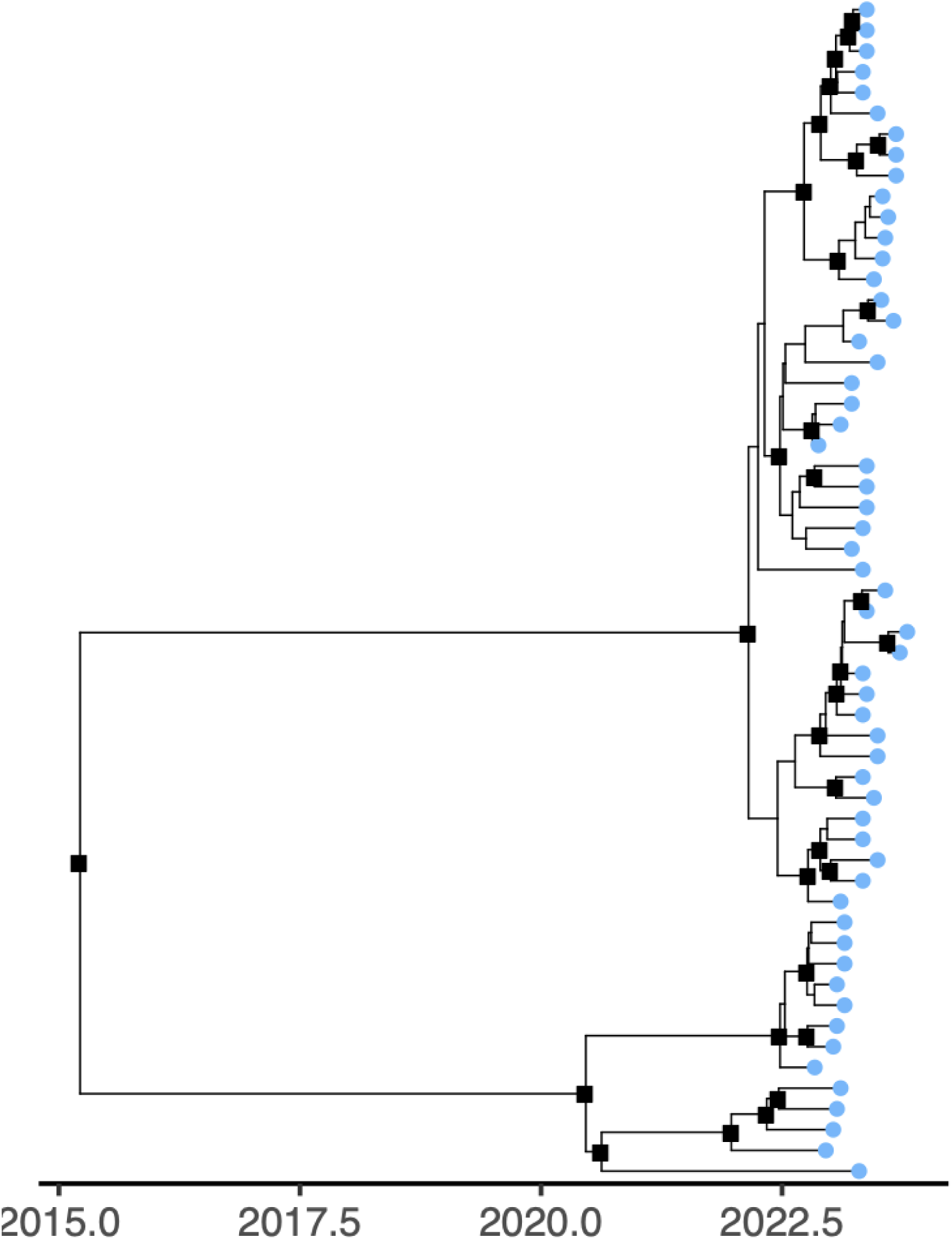
Time-scaled BEAST phylogenetic tree of *Klebsiella pneumoniae* (*Kpn*) sequence type 45. Blue tip color indicates colonization isolate, and posterior branch supports more than 0.75 are indicated with black boxes. Isolates were identified from colonization screening (skin or perirectal) for multidrug-resistant *Kpn* collected during surveillance; no detections from environmental or bloodstream infection isolates during this period.

**Supplementary Figure 2.**
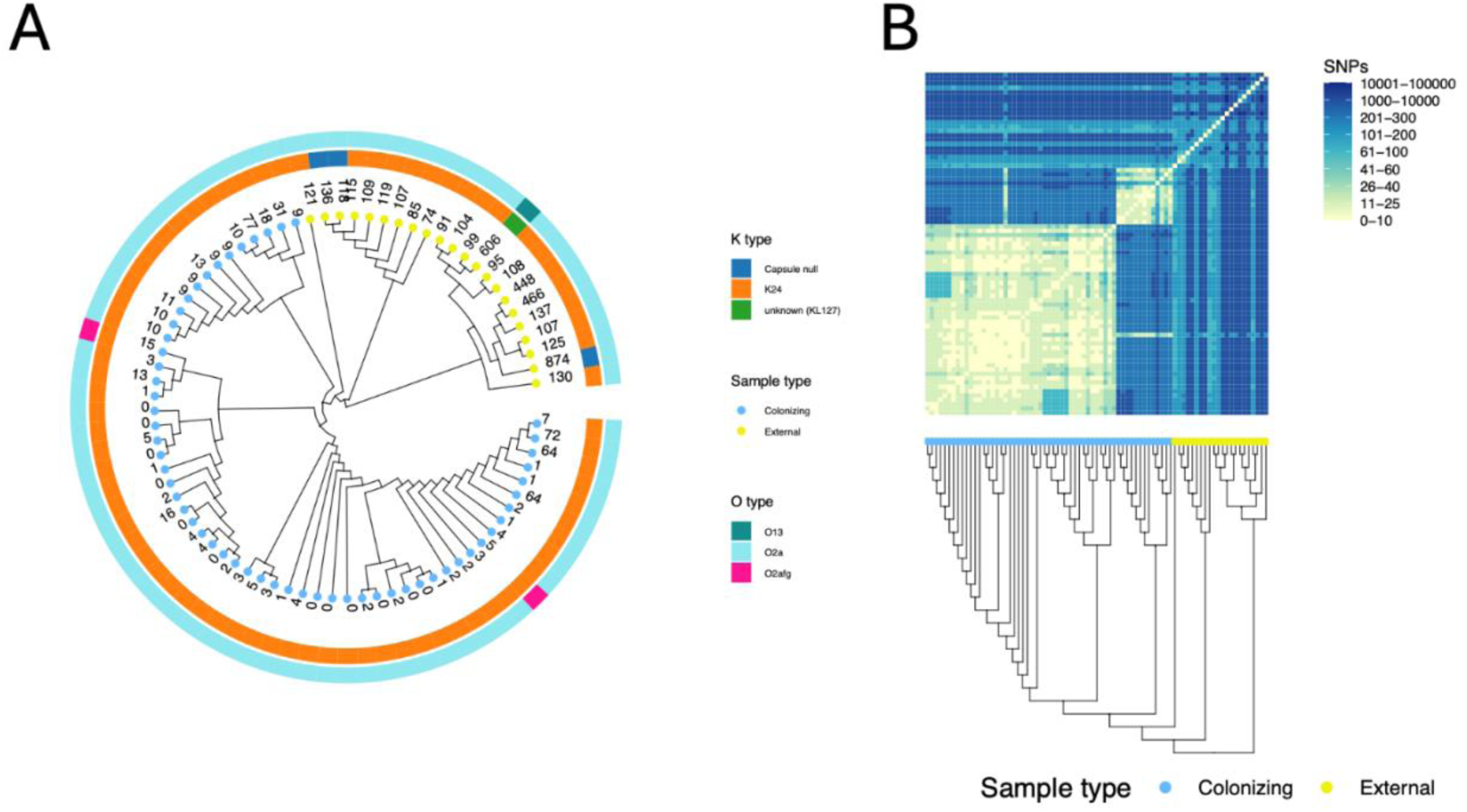
A. Maximum likelihood phylogenetic tree of ST45 *Klebsiella pneumoniae* isolates shown as a cladogram based on alignment to the best quality genome *SWEEP.APR5.19.05.23* within the ST using Snippy. Numbers on the tips represent the SNP distance from the reference. **B.** A heatmap produced using SNP-dist. Closest public genomes from NCBI were identified using WhatsGNU and were included. Blue and yellow tip color indicate study colonization isolates and external public genomes, respectively.

**Supplementary Figure 3.**
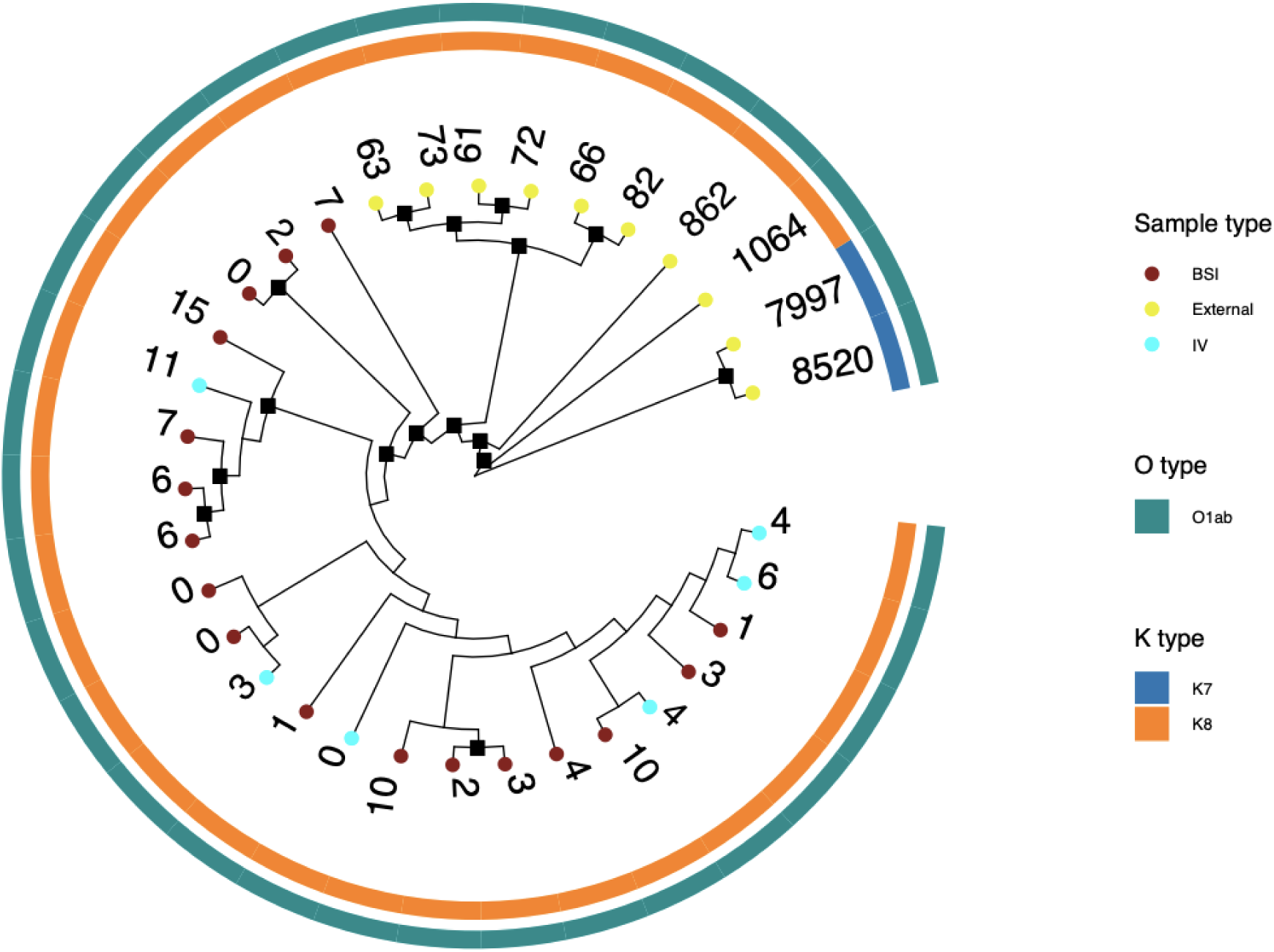
Maximum likelihood phylogenetic tree of ST1414 isolates shown as a cladogram based on alignment to the best quality genome ENV.NNU.28.dextrose.dup within the ST using Snippy. Closest public genomes from NCBI were identified using WhatsGNU and were included. Bootstrap values above 75 are shown on the branches as black squares. Numbers on the tips represent the SNP distance from the reference. Red, cyan and yellow tip colors indicate study bloodstream infection isolates, intravenous fluid isolates, and external public genomes, respectively.

**Supplementary Figure 4.**
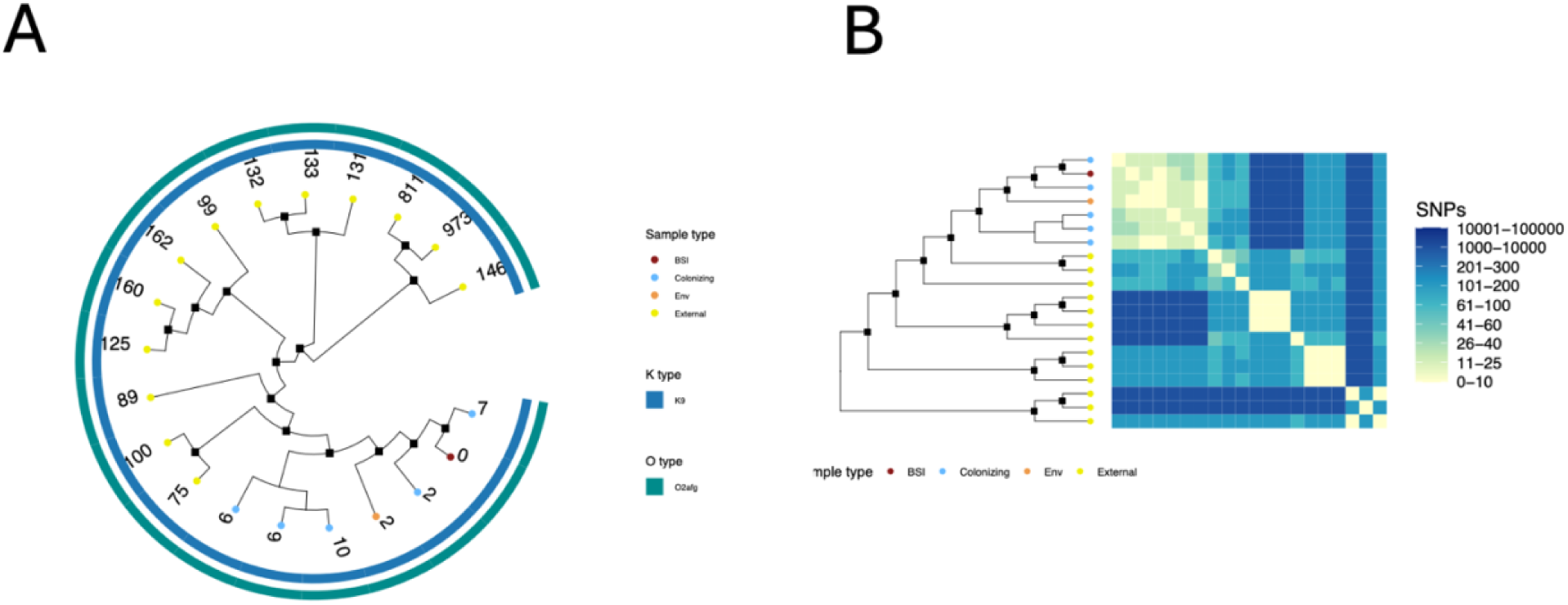
A. Maximum likelihood phylogenetic tree of ST22 isolates shown as a cladogram based on alignment to the best quality genome *CLIN.MF5126.dup* within the ST using Snippy. **B.** A heatmap produced using SNP-dist. Closest external public genomes from NCBI were identified using WhatsGNU and were included. Bootstrap values above 75 are shown on the branches as black squares. Numbers on the tips represent the SNP distance from the reference. Blue, red, orange, and yellow tip colors indicate study colonization isolates, bloodstream infection isolates, environmental isolates, and external public genomes, respectively.

**Supplementary Figure 5.**
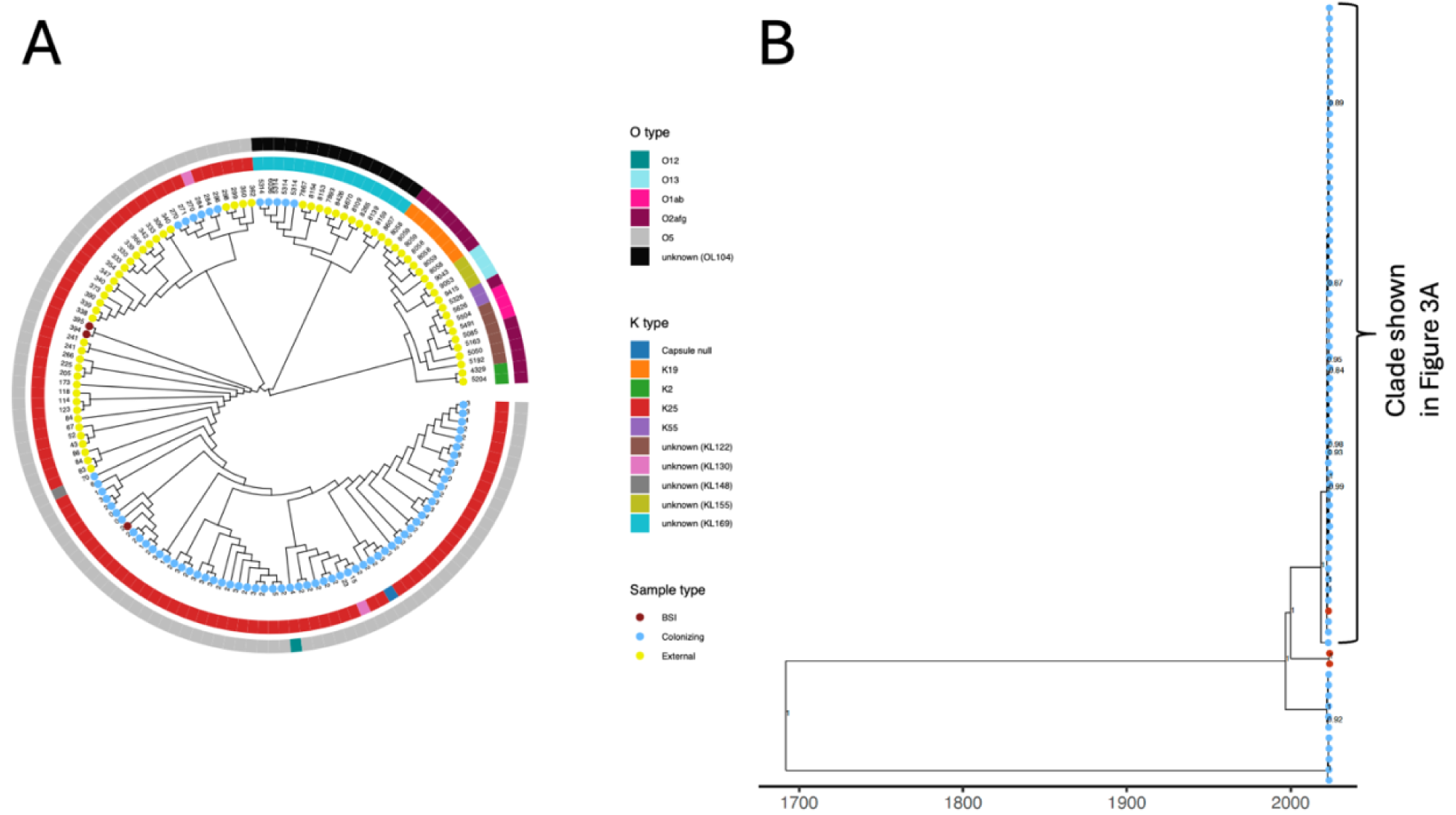
A. Maximum likelihood phylogenetic tree of ST17 isolates shown as a cladogram based on alignment to the best quality genome *SWEEP.PPS2.17.01.23* within the ST using Snippy. Closest public genomes from NCBI were identified using WhatsGNU and were included. Bootstrap values above 70 are shown on the branches. Numbers on the tips represent the SNP distance from the reference. Blue, red and yellow **t**ip color indicate colonization, bloodstream (BSI) isolates, and external public genomes, respectively. **B.** Time-scaled BEAST phylogenetic tree of *Klebsiella pneumoniae* ST17. Blue, red, and yellow tip colors indicate colonization isolates, BSI isolates, and external public genomes respectively. Posterior branch supports above 0.75 are indicated where relevant.

**Supplementary Figure 6.**
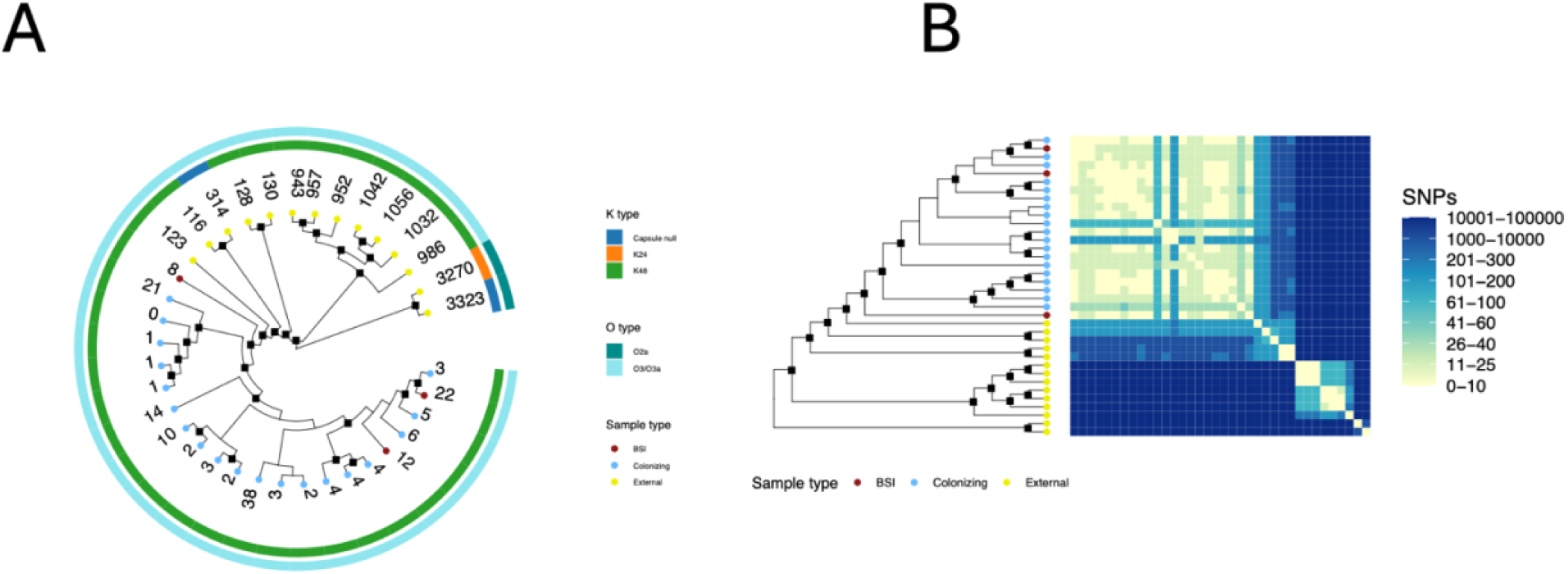
A. Maximum likelihood phylogenetic tree of S1805 isolates shown as a cladogram based on alignment to the best quality genome *SWEEP.FPR7.19.10.23* within the ST using Snippy. Numbers on the tips represent the SNP distance from the reference. Blue, red and yellow **t**ip color indicate colonization, bloodstream (BSI) isolates, and external public genomes, respectively. **B.** a heatmap produced using SNP-dist. Closest public genomes from NCBI were identified using WhatsGNU and were included. Bootstrap values above 75 are shown on the branches as black squares. Numbers on the tips represent the SNP distance from the reference. Blue, red, and yellow tip colors indicate colonization isolates, BSI isolates, and external public genomes respectively. Bootstrap values above 75 are shown on the branches as black squares.

**Supplementary Figure 7.**
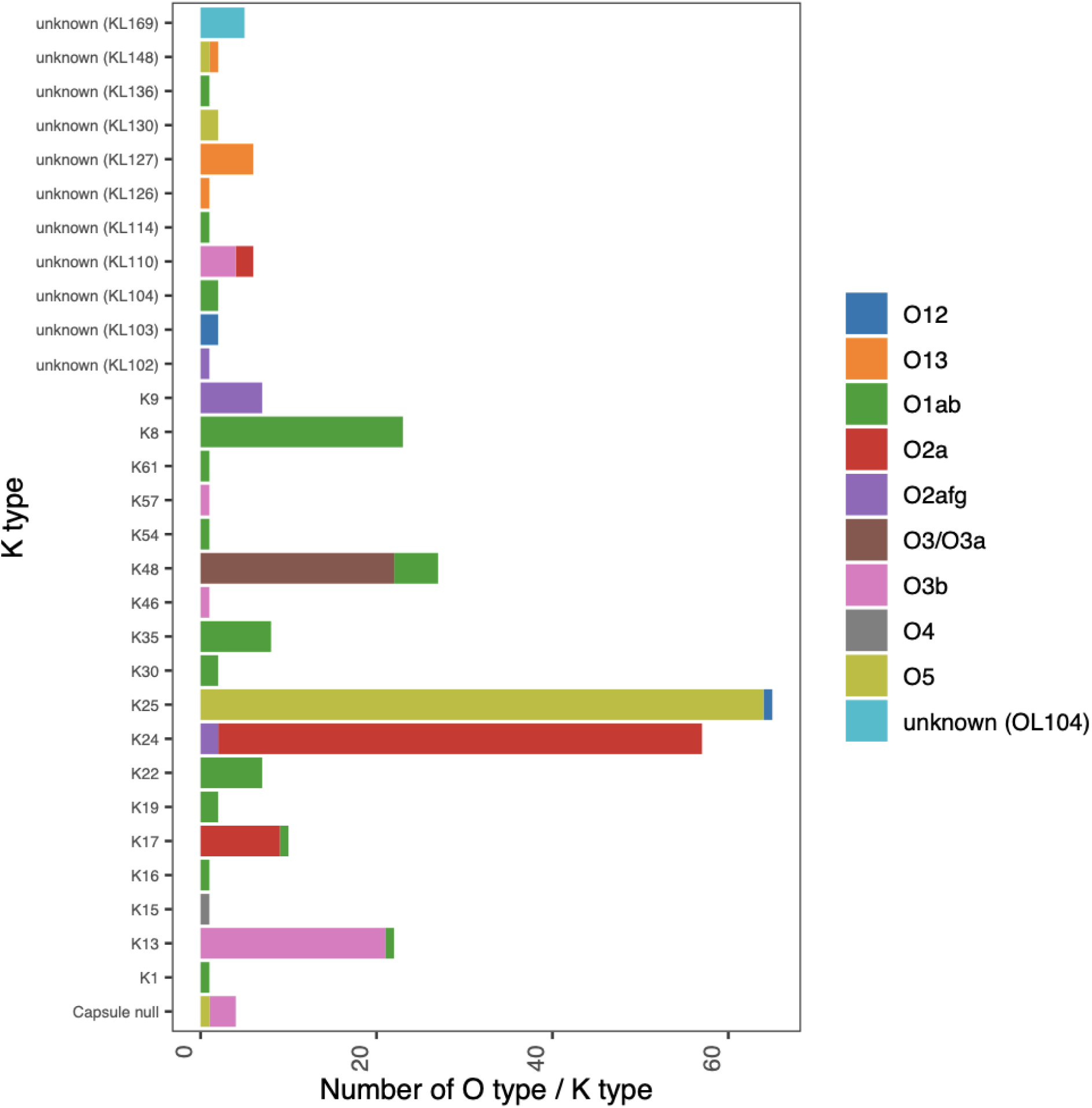
Distribution of K and O antigen types among *Klebsiella pneumoniae* isolates. Each bar represents the number of isolates with a given capsular (K) type, colored by associated O antigen type. Dominant K types include K25, K24, K48, K13 and K8.

